# Association between consumption of fermented vegetables and COVID-19 mortality at a country level in Europe

**DOI:** 10.1101/2020.07.06.20147025

**Authors:** Susana C Fonseca, Ioar Rivas, Dora Romaguera, Marcos Quijal, Wienczyslawa Czarlewski, Alain Vidal, Joao A Fonseca, Joan Ballester, Josep M Anto, Xavier Basagana, Luis M Cunha, Jean Bousquet

**Affiliations:** GreenUPorto - Sustainable Agrifood Production Research Centre, DGAOT, Faculty of Sciences, University of Porto, Campus de Vairão, Rua da Agrária, Porto, Portugal; ISGlobAL, Centre for Research in Environmental Epidemiology (CREAL), Barcelona, Spain; Medical Consulting Czarlewski, Levallois, France; MASK-air, Montpellier, France; World Business Council for Sustainable Development (WBCSD), Maison de la Paix, Geneva, Switzerland & AgroParisTech - Paris Institute of Technology for Life, Food and Environmental Sciences, Paris, France; CINTESIS, Center for Research in Health Technology and Information Systems, Faculdade de Medicina da Universidade do Porto; and Medida, Lda Porto, Portugal; IMIM (Hospital del Mar Research Institute), Barcelona, Spain; Universitat Pompeu Fabra (UPF), Barcelona, Spain; CIBER Epidemiología y Salud Pública (CIBERESP), Barcelona, Spain; Charité, Universitätsmedizin Berlin, Humboldt-Universität zu Berlin, and Berlin Institute of Health, Comprehensive Allergy Center, Department of Dermatology and Allergy, Berlin, Germany; MACVIA-France, Montpellier, France; Health Research Institute of the Balearic Islands (IdISBa), Palma de Mallorca, Spain; CIBER Physiopathology of Obesity and Nutrition (CIBEOBN), Madrid, Spain

**Author notes:** **Address for correspondance:** Professor Jean Bousquet, 273 avenue d’Occitanie, 34090 Montpellier, France.

**Keywords:** COVID-19, fermented foods, country, fermented vegetables, death rates

## Abstract

**Background:** Many foods have an antioxidant activity and nutrition may mitigate COVID-19. Some of the countries with a low COVID-19 mortality are those with a relatively high consumption of traditional fermented foods. To test the potential role of fermented foods in COVID-19 mortality in Europe, we performed an ecological study.

**Methods:** The European Food Safety Authority (EFSA) Comprehensive European Food Consumption Database was used to study the country consumption of fermented vegetables, pickled/marinated vegetables, fermented milk, yoghurt and fermented sour milk. We obtained the COVID-19 mortality per number of inhabitants from the Johns Hopkins Coronavirus Resource Center. EuroStat data were used for data on potential confounders at the country level including Gross Domestic Product (GDP) (2019), population density (2018), percentage of people older than 64 years (2019), unemployment rate (2019) and percentage obesity (2014, to avoid missing values). Mortality counts were analyzed with quasi-Poisson regression models - with log of population as an offset - to model the death rate while accounting for over-dispersion.

**Results:** Of all the variables considered, including confounders, only fermented vegetables reached statistical significance with the COVID-19 death rate per country. For each g/day increase in the average national consumption of fermented vegetables, the mortality risk for COVID-19 decreased by 35.4% (95% CI: 11.4%, 35.5%). Adjustment did not change the point estimate and results were still significant.

**Discussion:** The negative ecological association between COVID-19 mortality and consumption of fermented vegetables supports the *a priory* hypothesis previously reported. The hypothesis needs to be tested in individual studies performed in countries where the consumption of fermented vegetables is common.

## Introduction

One of the striking problems raised by the COVID-19 pandemic is the highly variable death rate between and within countries. In Europe, countries like Belgium, France, Italy, Spain and the UK have a very high death rate, whereas central European countries, the Balkans and some Nordic countries have very low rates (1). Similar differences exist globally. In Europe, some countries with low mortality rates, like Germany, initiated early tests and lockdown (2). However, this is unlikely to be the only explanation. The COVID-19 epidemic is multifactorial. Factors like climate, population density, age structure, socio-economic factors (such as unemployment, but also housing conditions, including those at retirement homes) are also associated with increased incidence and mortality.

Among other potentially relevant factors, diet has received little attention. Many foods have an antioxidant activity (3-5) and it has been proposed that the role of nutrition may mitigate COVID-19 (6). Some processes like fermentation increase the antioxidant activity of milk, cereals, fruit, vegetables, meat and fish (7). Some countries with low COVID-19 mortality rates appear to be those with a relatively high consumption of traditional fermented foods (1).

To test the potential role of fermented foods in the COVID-19 mortality in Europe, we have performed an ecological study.

## Methods

### Selection of the database

The European Union provided the European Food Safety Authority (EFSA) Comprehensive European Food Consumption Database in 2011 as well as an update in 2015. This contains detailed data for EU countries (https://www.efsa.europa.eu/en/food-consumption/comprehensive-database). It enables the study of the assessment of nutrient intake in the populations of many EU countries. In the database, dietary surveys and food consumption data for each country are divided into categories. These include: age (from infants to adults aged 75 years or older); food group (over 2,500) and type of consumption (covering both regular and high consumption).

The Comprehensive Database constitutes a unique resource for the estimation of consumption figures across the European Union and represents a useful tool for assessing nutrient intake in Europe. However, many substantial methodological differences exist between countries, data are provided over a rather long period of time and data should be considered with caution (8). To generate maps with the distribution of consumption, we used 25, 50 and 75% confidence intervals (CI) to classify the countries’ food intake (low <25%, medium: 25-49%, high: 50-75%, very high: >75%) in adults.

Since data from certain countries are missing in the database, we attempted to find other sources of data. We obtained data from Poland for fermented foods (Polish Academy of Science) (9).

### Selection of foods

The hypothesis raised recently proposed that diet with traditional fermented foods may explain some of the differences in the COVID-19 death rates between countries (1). We then selected fermented foods from the database that are not necessarily prepared using traditional recipes (“fermented milk products”) and others that are more likely to be closer to traditional foods (“fermented vegetables”, and “traditional sour milk”). We added “probiotic milk”, “yoghurt” and “pickled/marinated vegetables” to this list (Table 1).

**Table 1:** Foods tested.

| Exposure hierarchy<br>(L1) | Exposure hierarchy<br>(L2) | Exposure hierarchy<br>(L3) | Exposure hierarchy<br>(L4) | N<br>countries | N<br>subjects |
| --- | --- | --- | --- | --- | --- |
| Milk and dairy<br>products | Fermented milk or<br>cream | Fermented milk<br>products | Traditional sour milk<br>products | 17 | 33,279 |
|  |  |  | Probiotic milk-like<br>drinks | 9 | 16,474 |
|  |  |  | Yoghurt | 22 | 40,143 |
| Vegetables and<br>vegetable products | Processed or<br>preserved vegetables<br>and similar | Fermented or<br>pickled vegetables | Fermented vegetables | 17 (+1) | 33,816 |
|  |  |  | Pickled vegetables | 17 | 33,749 |

For fermented vegetables, Polish data were obtained from other sources

Data from the national surveys of different EU countries have been extracted from the EFSA Comprehensive European Food Consumption Database for different fermented foods (Table 2).

**Table 2:** Origin (survey and year) of consumption data obtained from the EFSA Comprehensive European Food Consumption database.

|  |  |  | Fermented<br>vegetables | Fermented<br>sour milk | Yoghurt | Probiotic<br>milk | Marinated<br>vegetables |
| --- | --- | --- | --- | --- | --- | --- | --- |
| Austria | 2014 | AT-NATIONAL-2016 | + | + | + |  | + |
| Belgium | 2014 | NATIONAL-FCS-2014 | + | + | + | + | + |
| Croatia | 2011 | NIPNOP-HAH-2011-2012 | + | + | + | + | + |
| Czech Republic | 2003 | SISP04 | + | + | + | + | + |
| Denmark | 2013 | DANSDA 2005-08 |  |  | + | + |  |
| Estonia | 2013 | DIET-2014-EST-A | + | + | + | + | + |
| Finland | 2012 | FINDIET2012 | + | + | + | + |  |
| France | 2014 | INCA3 | + | + | + | + | + |
| Germany | 2007 | NATIONAL NUTRITION SURVEY II | + | + | + |  |  |
| Greece | 2014 | GR-EFSA-LOT2 2014-2015 |  | + | + |  | + |
| Hungary | 2003 | NATIONAL REPR SURV | + |  | + |  | + |
| Ireland | 2008 | NANS 2012 | + | + | + |  |  |
| Italy | 2005 | INRAN SCAI 2005-06 | + |  | + |  | + |
| Latvia | 2012 | LATVIA_2014 | + | + | + |  | + |
| Netherlands | 2012 | FCS2016_CORE | + | + | + | + | + |
| Portugal | 2015 | IAN.AF 2015-2016 | + | + | + | + | + |
| Romania | 2012 | DIETA PILOT ADULTS | + | + | + |  | + |
| Slovenia | 2017 | SI.MENU-2018 | + | + | + |  | + |
| Spain | 2013 | ENALIA2 | + | + | + | + | + |
| Sweden | 2010 | RIKSMATEN 2010 | + | + | + | + | + |
| United Kingdom | 2008 | NDNS ROLLING PROGRAMME YEARS 1-3 | + |  | + | + |  |

### Assessment of COVID-19 mortality

We used mortality per number of inhabitants to assess the death rates as proposed by the European Center for Disease Prevention and Control (ecdc, https://www.ecdc.europa.eu/en). To create maps with the death rate distribution, we used cutoffs at 25 (very low death rates), 50 (low), 100 (medium), 250 (high) and 500 (very high) per million (May 20, 2020) (1). Raw data were used for the Poisson analysis.

Data on mortality for COVID-19 were downloaded from the Johns Hopkins Coronavirus Resource Center (https://coronavirus.jhu.edu) on June 22^nd^, 2020. In addition, we downloaded data from EuroStat on: Gross Domestic Product (GDP) of the country (2019), population density (2018), percentage of people older than 64 years (2019), unemployment rate (2019) and percentage of obesity (2014, to avoid missing values).

### Statistical analysis

Mortality counts were analyzed with quasi-Poisson regression models - with log of population as an offset - to model the death rate while accounting for over-dispersion. Models were fitted separately for each of the food consumption groups. Due to the small number of countries included in the analyses, we adjusted our models using the first principal component of a principal component analysis (PCA) of the variables GDP, population density, percentage of people older than 64%, unemployment and obesity (10).

## Results

### Death rates in European countries

The death rate per million people in the European Union varies widely. There is a gradient between Western countries with a high death rate and those North of the Alps or Balkans with a low (50-100 deaths per million) or very low (under 50/million) death rate. One exception is Sweden with a high death rate (Figure 1).

**Figure 1:**
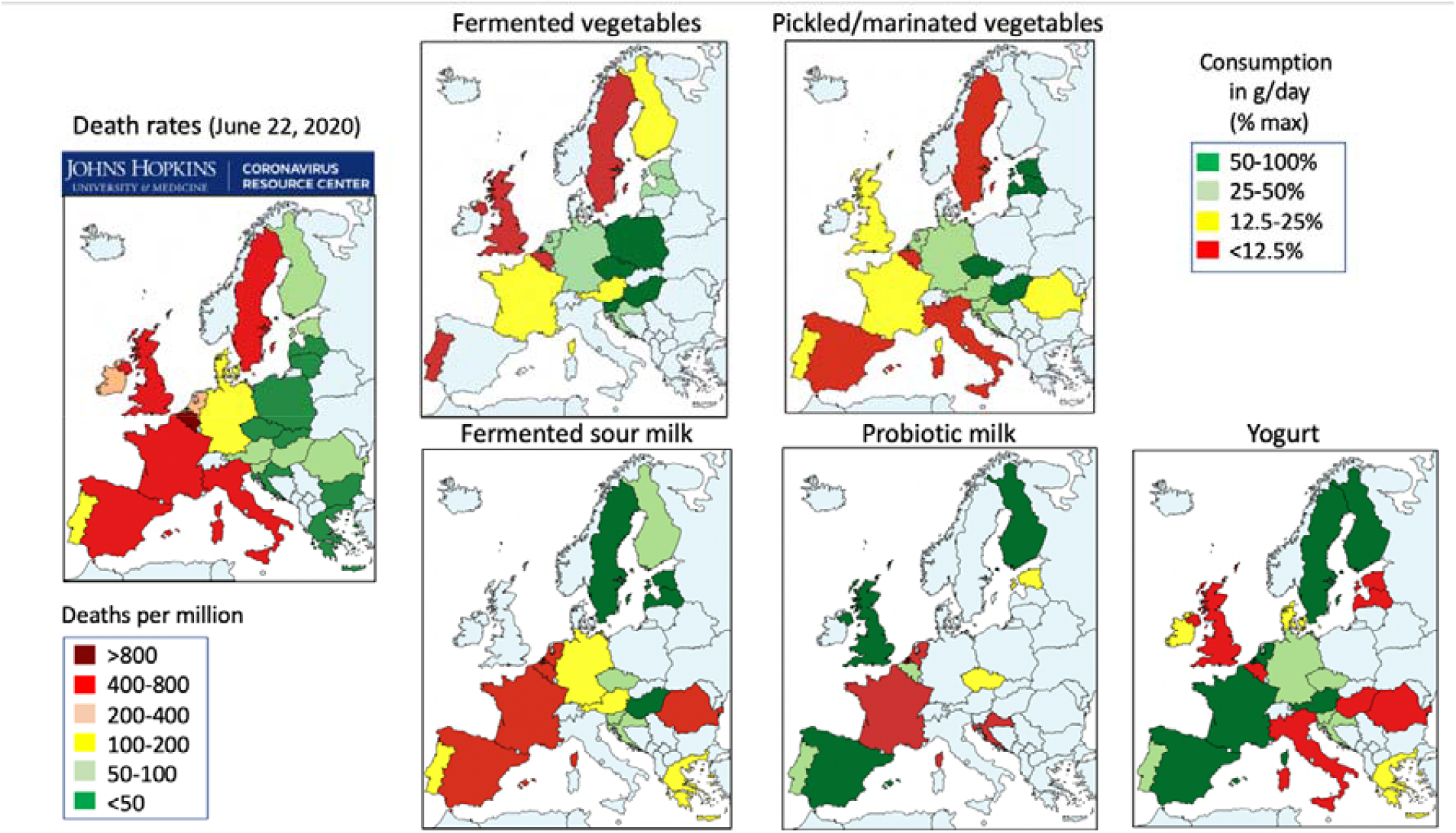
COVID-19 death rate and consumption of foods in European Union countries.

### Food intake in European countries

The intake of selected foods was given in g/day (Figure 1). Yoghurt and fermented milk are widely consumed in most EU countries with no apparent difference between low and high-death rate countries. Many countries have a low consumption of fermented sour milk. Only 4/17 countries have a relatively high consumption and three have a low death rate. Sweden has a high consumption with a high death rate. There are few data for probiotic milk but there appears to be no association with death rates.

Fermented vegetables are mostly consumed in low death rate countries. Few countries consume pickled/marinated vegetables but countries with a consumption of these foods have a low death rate.

### Association between COVID-19 mortality and food intake at the country level

Of all the variables considered, including confounders, only fermented vegetables reached statistical significance (unemployment was close to significance, with p-value 0.06) with COVID-19 death rate per country (Table 3 and Figure 2).

**Table 3.**
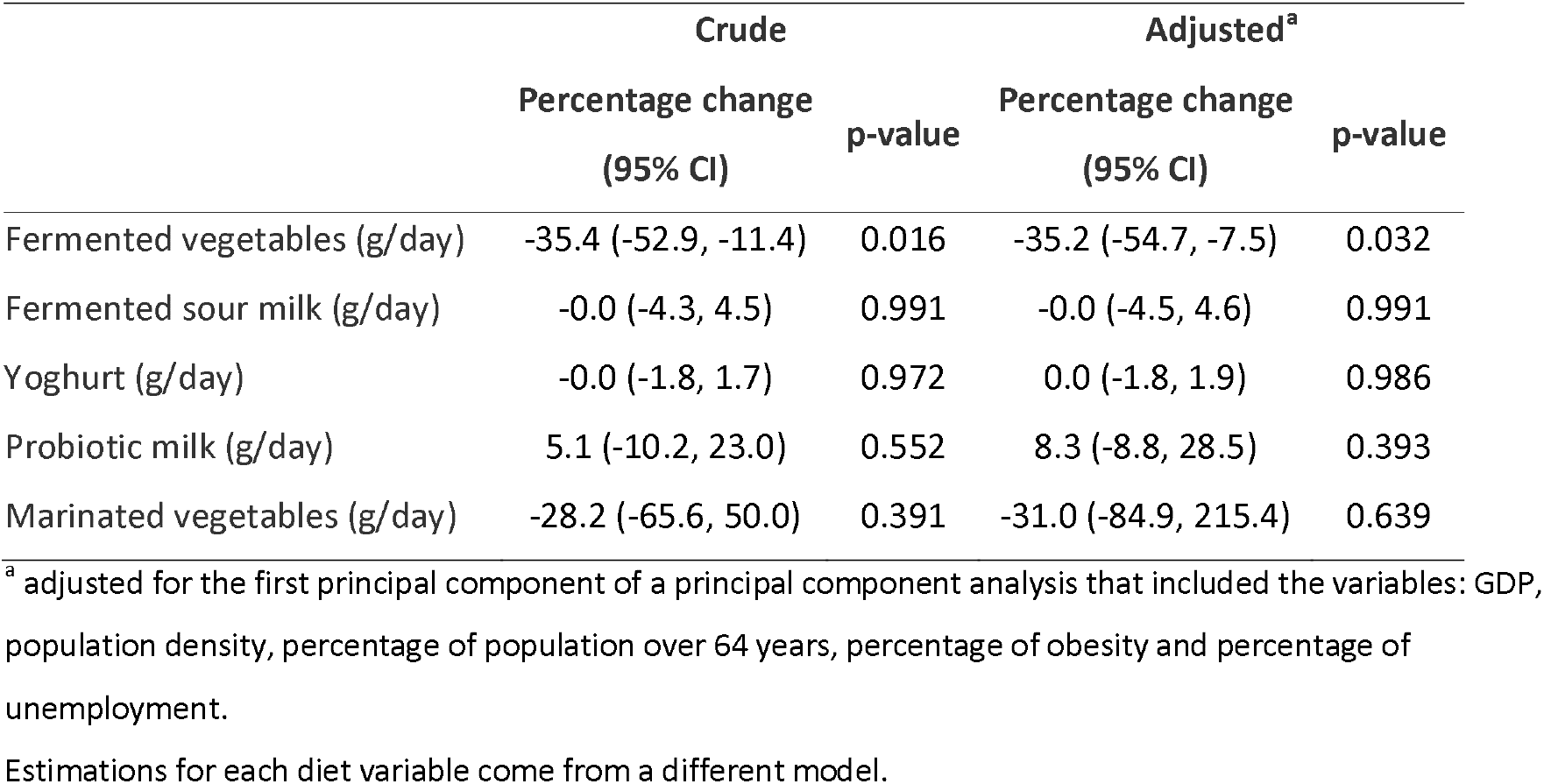
Percentage change in COVID-19 death risk (with 95% confidence intervals (CI)) as a function of food group consumption.

|  | Crude |  | Adjusted<sup>a</sup> |  |
| --- | --- | --- | --- | --- |
|  | Percentage change<br>(95% CI) | p-value | Percentage change<br>(95% CI) | p-value |
| Fermented vegetables (g/day) | -35.4 (-52.9, -11.4) | 0.016 | -35.2 (-54.7, -7.5) | 0.032 |
| Fermented sour milk (g/day) | -0.0 (-4.3, 4.5) | 0.991 | -0.0 (-4.5, 4.6) | 0.991 |
| Yoghurt (g/day) | -0.0 (-1.8, 1.7) | 0.972 | 0.0 (-1.8, 1.9) | 0.986 |
| Probiotic milk (g/day) | 5.1 (-10.2, 23.0) | 0.552 | 8.3 (-8.8, 28.5) | 0.393 |
| Marinated vegetables (g/day) | -28.2 (-65.6, 50.0) | 0.391 | -31.0 (-84.9, 215.4) | 0.639 |

<sup>a</sup> adjusted for the first principal component of a principal component analysis that included the variables: GDP, population density, percentage of population over 64 years, percentage of obesity and percentage of unemployment.
Estimations for each diet variable come from a different model.

**Figure 2.**
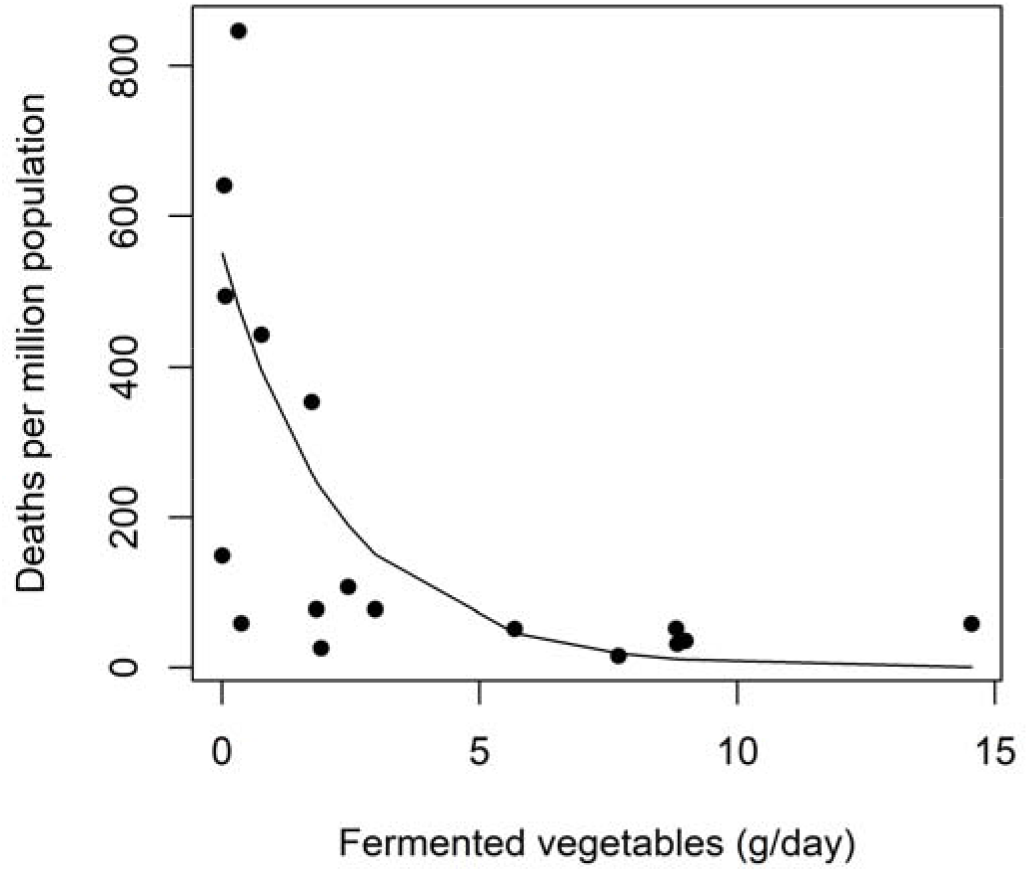
Raw data and estimated relationship between country death rates for COVID-19 and consumption of fermented vegetables (g/day) using the quasi-Poisson regression model.

For each g/day increase in consumption of fermented vegetables of the country, the mortality risk for COVID-19 decreased by 35.4% (CI 95% −54.7, −7.5). Adjustment for the first principal component of the confounders did not change the point estimate but it widened the confidence intervals, although the association was still significant. The association with fermented vegetables was also significant if the model was adjusted for unemployment instead of the first principal component.

## Discussion

This study shows that in countries where fermented vegetables are highly consumed, death rates are low. On the other hand, there was no statistical significance on percentage change (p-value > 0.05) with pickled/marinated vegetables or probiotic milk products and no correlation (percentage change zero) with fermented sour milk and yoghurt.

### Limitations

According to the Johns Hopkins coronavirus resource center (https://coronavirus.jhu.edu), one of the most important ways of measuring the burden of COVID-19 is mortality. However, death rates are assessed differently between countries and there are many biases that are almost impossible to control (1). Using the rate of COVID-19 confirmed cases is subject to limitations. Differences in the mortality rates depend on the characteristics of the health care system, the reporting method, whether or not deaths outside the hospital have been counted and other factors, many of which remain unknown. Countries throughout the world have reported very different case fatality ratios - the number of deaths divided by the number of confirmed cases - but these numbers cannot be compared easily due to biases.

It is very important to consider differences in food consumption within countries but this cannot be studied using the EFSA database. As found in France, Spain and Italy, there are large regional differences in death rates and it would be of interest to compare sub-national regions with the different consumptions of fermented foods (1).

A limited number of countries have been studied due to the lack of information on food consumption, and a definite conclusion cannot be made. The selection of confounding factors has been arbitrary and more are needed. As in any ecological study, any inference from the observed association should be made at the country level, as the possibility of ecological fallacy precludes inferences at the individual level. Further testing in properly designed individual studies would be of interest.

Moreover, associations do not mean causality, and patterns of food consumption can be a proxy for other undetermined factors.

Understanding the multiple factors that are likely to explain inter and intra-national geographical variations will not be possible until indicators of the pandemic distribution have been improved and properly-designed studies conducted. However, this study is of interest since it is the first to attempt to link death rates with food consumption at a country level.

### Interpretation

COVID-19, like most diseases, exhibits large geographical variations which frequently remain unexplained despite abundant research (11). Though the more relevant factors are likely to be related to intensity and timing of interventions (12), demographic factors, seasonal variations, immunity, and cross-immunity, other factors like environment or nutrition should not be overlooked. Our study suggests that European countries with a lower COVID-19 mortality were more likely to exhibit a large consumption of fermented foods.

Though our results don’t allow to infer causality, they do reinforce our *a priory* hypothesis that the ingestion of fermented food may have reduced the severity of the COVID-19 due to its antioxidant activity. Many foods have an antioxidant activity (3-5). Several foods can interact with transcription factors related to antioxidant effects such as Nrf-2, (13). “Ancient foods”, and particularly those containing *Lactobacillus*, activate Nrf2 (14). Effects of microbiome (15) and diet (1, 16) on SARS-CoV-2 infection suggest that their role in the gut could be a target for COVID-19 interventions. Some patients with COVID-19 have intestinal microbial dysbiosis with decreased probiotics such as *Lactobacillus* and *Bifidobacterium* (17).

Our hypothesis is biologically plausible. Mechanisms underlying these results may be approached. The increased severity of COVID-19 in diabetes, hypertension, obese or old age individuals may be related to insulin resistance with oxidative stress as a common pathway (18, 19). SARS-CoV-2 binds to the angiotensin converting enzyme II (ACE-2). The ACE-2 receptor is a part of the dual system – the renin-angiotensin-system (RAS) - consisting of an ACE-Angiotensin-II-AT_1_R axis and an ACE-2-Angiotensin-(1-7)-Mas axis. The AT_1_R axis is involved in most of the effects of Ang II, such as oxidative stress generation (20). In metabolic disorders and with increased age, there is an upregulation of the AT_1_R axis leading to pro-inflammatory and pro-fibrotic effects in the respiratory system (21). The Nrf2 (nuclear factor erythroid 2 p45-related factor 2), a transcription factor that protects against oxidative stress, may be involved in diseases associated with insulin-resistance (22-24). Nrf2 activation may decrease the intensity of the cytokine storm seen in COVID-19 (25).

Against our expectations, we did not find an association between COVID-19 mortality and consumption of fermented milk. Milk products also contain LABs (26, 27) but their protective effect in diseases associated with insulin resistance is not easy to demonstrate. A large meta-analysis found that the intake of probiotics resulted in minor but consistent improvements in several metabolic risk factors in subjects with metabolic diseases, and particularly in insulin resistance (28). More studies are needed to test whether other types of fermented foods show an association with COVID-19 severity.

The results of the study are of interest since we found that fermented milk is not associated with death rates, in contradiction to traditional foods as was earlier proposed (1). This finding may be of importance for the optimization of processed fermented foods in order to obtain either a spectrum of LABs closer to traditional fermented foods and/or a level of daily consumption required to be protective.

Our study was restricted to European countries, so its results cannot be extrapolated to other regions. However, other regions with low COVID-19 mortality also exhibit a large consumption of fermented foods. In Asia, where the pandemia started, death rates are very low and the pandemia appears to be under control. The same happened in Africa where the COVID-19 spread was predicted to be catastrophic and death rates appear to be low. The climate and the low population density (as in rural areas but not in megacities such as Cairo or Lagos) or the differences in health infrastructure are unlikely to be the only explanation (29, 30). In Asia, fermented vegetables are highly consumed (31-34). In Africa, a major food is manioc, usually obtained by the fermentation of cassava tubers (35). It would be of great importance to confirm these data using food consumption in Asian countries and to perform definitive epidemiologic and mechanistic studies to confirm the present paper.

If the hypothesis is proved, COVID-19 will be the first infectious disease epidemic whose biological mechanisms are proved to be associated with a loss of “nature” (36). Imbalance in the gut microbiota is associated with the pathogenesis of various disease types including allergy, asthma, rheumatoid arthritis, different types of cancer, diabetes mellitus, obesity and cardiovascular disease (37). Fermentation was introduced during the Neolithic age and was essential for the survival of human kind. When modern life led to eating reduced amounts of fermented foods, the microbiome drastically changed (38) and this may have facilitated SARS-CoV-2 to spread or to be more severe (39). The latter would add to other mechanisms linking the COVID-19 with the ecological disruption of the Anthropocene (40).

Thus, although this study is only indicative of the role of diet in COVID-19, it is however another piece of the hypothesis proposing that traditional fermented foods may be involved in the prevention of severe COVID-19 at a country level.

## Data Availability

The data are available on request to Prof Jean Bousquet

## Acknoledgment

We would like to thank Manuel Moñino for his help with and advice on the use of the EFSA data

